# Prevalence, Awareness and Correlates of Self-Reported Hypoglycemia Among Adults with Type 2 Diabetes in Northern Jordan: A Cross-Sectional Study

**DOI:** 10.64898/2026.10.29.26366815

**Authors:** Adi Khasawneh, Taima’a Kefah Al-Mohammad, Abdalrahman Abu Qutaish, Karam Mohamad Aldeeb, Sohaib Fawaz Shaheen, Razan Yaseen, Sara Ali Nassar, Mohammad Emad Melhem, Abdel-Hameed Al-Mistarehi, Heba Okour, Khalid A. Kheirallah

**Author notes:** Corresponding author Khalid A. Kheirallah, Department of Public Health and Family Medicine, Jordan University of Science and Technology, P.O. Box 3030, Irbid 22110, Jordan. E-mail: [ ].

## Abstract

Hypoglycemia is a frequent and potentially dangerous adverse effect of glucose-lowering therapy in type 2 diabetes mellitus (T2DM), yet data on its prevalence, patient awareness and correlates in Jordan are limited. We conducted a cross-sectional study of 326 adults with T2DM attending family medicine outpatient clinics at a tertiary teaching hospital (King Abdullah University Hospital, KAUH) and a university-affiliated primary healthcare center (JUST PHC) in northern Jordan between 2025 and 2026. Trained interviewers administered a structured questionnaire, and laboratory results were verified from electronic medical records. Hypoglycemia was defined as a glucose level below 70 mg/dL or symptoms relieved by fast-acting carbohydrate during the preceding six months. Prevalence was estimated with 95% confidence intervals, and associated factors were examined using chi-square tests and multivariable logistic regression. The prevalence of self-reported hypoglycemia was 79.1% (258/326; 95% CI 74.4-83.2). Although 71.5% of participants knew the symptoms of hypoglycemia, 73.9% were unaware that it can occur without symptoms, 53.6% had never reported an episode to a healthcare provider, only 31.3% had discussed hypoglycemia risk with a physician, and 50.6% never carried sugar or an emergency kit. In the multivariable model, insulin use (adjusted odds ratio [aOR] 3.27, 95% CI 1.75-6.10), sulfonylurea use (aOR 2.14, 1.18-3.87), diabetic nephropathy (aOR 3.76, 1.05-13.48) and attending the primary healthcare center versus the hospital (aOR 2.76, 1.42-5.36) were independently associated with higher odds of hypoglycemia, whereas beta-blocker use was associated with lower odds (aOR 0.52, 0.28-0.95). Self-reported hypoglycemia was highly prevalent among patients with T2DM in northern Jordan, and awareness of asymptomatic hypoglycemia and reporting to providers were limited. Routine structured enquiry about hypoglycemia, patient education that addresses asymptomatic episodes, and review of high-risk therapy, particularly in primary care, are warranted.

## Introduction

Diabetes mellitus (DM) represents a major and growing global health challenge, with substantial morbidity, mortality, and economic burden. In Jordan, the estimated prevalence of diabetes among adults reached 20.5% in 2024, reflecting a significant national public health concern [1]. Achieving and maintaining optimal glycemic control remains central to preventing long-term complications; however, hypoglycemia constitutes a major barrier to attaining recommended glycemic targets, particularly when intensive glycemic control strategies are pursued [2–4].

In individuals with DM, hypoglycemia is defined as any abnormally low plasma glucose concentration that may cause harm, regardless of the presence or severity of symptoms [3,4]. Multiple risk factors have been associated with hypoglycemia, including advanced age; longer duration of diabetes; insulin or sulfonylurea use; irregular meal patterns; fasting; comorbid conditions such as renal impairment; and prior hypoglycemic episodes; particularly recurrent events that contribute to impaired counter-regulation [2,5,6]. Beyond its immediate clinical manifestations, hypoglycemia carries significant long-term consequences. It has been linked to increased risk of cognitive impairment, cardiovascular events, and all-cause mortality [7–9]. In addition to its clinical burden, hypoglycemia imposes substantial strain on healthcare systems through emergency visits, hospitalizations, and indirect costs [10–13]. Recurrent episodes may also lead to impaired awareness of hypoglycemia due to autonomic failure, thereby increasing the risk of severe events [14–16].

Although hypoglycemia occurs more frequently in individuals with type 1 diabetes mellitus (T1DM), it also represents a substantial burden among patients with type 2 diabetes mellitus (T2DM). A meta-analysis of 46 population-based studies reported a pooled prevalence of 45% for mild-to-moderate and 6% for severe hypoglycemia among patients with T2DM, rising to 50% and 21%, respectively, among those treated with insulin [17]; risk is also higher with progressive pancreatic β-cell failure [2,3,6]. In Jordan, a study conducted at the National Center for Diabetes, Endocrinology and Genetics (NCDEG) reported that 67.7% of patients with T2DM were receiving insulin therapy, and 46.6% experienced hypoglycemia [18]. These findings underscore the magnitude of the problem in patients with T2DM receiving oral and injectable antihyperglycemic drugs as well as insulin therapy at the local setting.

Previous studies conducted in Jordan have examined the prevalence of hypoglycemia among patients with T2DM [18,19], including work restricted to elderly outpatients (≥60 years) at a single tertiary hospital in northern Jordan, focused primarily on clinical and pharmacological predictors of hypoglycemic episodes and their downstream healthcare utilization [19]. However, no local study to date has examined patients’ awareness of hypoglycemia-related signs and symptoms, distinguished reported from unreported episodes, or explored patients’ reasons for non-disclosure to their physicians, nor has any study compared hypoglycemia prevalence and reporting behavior across different family medicine outpatient settings (tertiary hospital-based versus non-tertiary, university-affiliated primary healthcare center) in Jordan.

This study aimed primarily to estimate the prevalence of self-reported hypoglycemia among patients with type 2 diabetes mellitus attending outpatient clinics in northern Jordan. Secondary objectives included evaluating patients’ awareness of hypoglycemia-related signs and symptoms, including awareness that hypoglycemia can occur without symptoms and their management; assessing blood glucose monitoring practices; and identifying risk factors associated with experiencing hypoglycemia.

## Methods

### Study Design and Settings

This was a cross-sectional study conducted in Family Medicine outpatient clinics at two sites in Irbid, Northern Jordan: King Abdullah University Hospital (KAUH), a tertiary care teaching hospital, and the Primary Healthcare Center of Jordan University of Science and Technology (JUST PHC), a non-tertiary, university-affiliated primary healthcare facility. Data were collected over a one-year period from April 2025 to January 2026, using a pretested questionnaire and electronic medical records to verify patient laboratory results.

### Ethical Considerations

This study was conducted in accordance with the Declaration of Helsinki and approved by the Institutional Review Board (IRB) of Jordan University of Science and Technology (JUST), which provides ethical oversight for research conducted at King Abdullah University Hospital (KAUH) and the JUST Primary Healthcare Center (JUST PHC). Approval was granted on 17 March 2025 (IRB reference number: Feb2025/179-30). Written informed consent was obtained from all participants, and participation was voluntary.

### Eligibility Criteria

The inclusion criteria were adults aged 18 years or older with a diagnosis of T2DM. The exclusion criteria were type 1 diabetes mellitus, a T2DM diagnosis of less than 6 months’ duration, and pregnancy.

### Recruitment and Data Collection

Patients with T2DM attending the Family Medicine outpatient clinics at KAUH and JUST PHC who met the inclusion criteria were invited to participate in this study. Of 355 eligible patients approached, 326 agreed to participate, yielding a response rate of 91.8%. After explaining the study objectives and obtaining informed consent, data collection was conducted during face-to-face interviews with patients by a team of trained interviewers. The electronic medical records were used to verify the most recent results of required laboratory tests. Study site (KAUH or JUST PHC) was derived from the medical file number prefix (KAUH-issued versus JUST/MOH-issued records).

### Questionnaire and Variables

The questionnaire was developed based on available guidelines and an extensive literature review, then reviewed by family physicians, epidemiologists, and public health experts. A pilot study was subsequently conducted on 52 participants from the Family Medicine outpatient clinics at KAUH and JUST PHC to test its feasibility.

The questionnaire included questions on socio-demographics; medical history related to DM, including the use of antihyperglycemic drugs, insulin type, and total dose; comorbidities; hypoglycemia; and the type and frequency of blood glucose monitoring, in addition to questions about patients’ follow-up laboratory tests and their most recent results. Medical records were also abstracted to validate self-reported DM history and laboratory testing.

Some questions assessed patients’ level of knowledge and awareness regarding asymptomatic hypoglycemia and hypoglycemia-related signs and symptoms. To ensure correct understanding and consistency, common signs and symptoms of hypoglycemia were explained to participants.

Patients were then asked whether they had experienced any hypoglycemic episodes during the past 6 months. Hypoglycemia was defined as either a measured blood glucose level below 70 mg/dL or self-reported symptoms consistent with hypoglycemia that resolved after ingestion of fast-acting carbohydrates. Symptoms of hypoglycemia include, but are not limited to, shakiness, irritability, confusion, tachycardia, dizziness, sweating, and hunger [20]. A hypoglycemic episode requiring an emergency department visit or hospital admission was classified as severe. Participants who met the criteria for hypoglycemia were asked about the frequency, time of occurrence, and management of the event. Participants were also asked whether anxiety or fear related to hypoglycemia had affected their adherence to antihyperglycemic drugs, whether they had ever taken an incorrect medication dosage, about unreported hypoglycemia and reasons for non-disclosure, and whether they had received verbal or written instructions from their healthcare provider regarding hypoglycemia management.

### Statistical Analysis

Data were analyzed using IBM SPSS Statistics version 26. Categorical variables were summarized as frequencies and percentages, and the prevalence of hypoglycemia is presented with Wilson score 95% confidence intervals. Unadjusted associations between patient characteristics and self-reported hypoglycemia were assessed using Pearson’s chi-square test (with continuity correction for 2×2 tables). Prior to multivariable modeling, multicollinearity was assessed, and variables showing severe collinearity or confounding by indication were purposefully excluded; diabetic nephropathy and chronic kidney disease overlapped almost completely (21 of 22 patients with CKD also had nephropathy), so only nephropathy, the diabetes-specific measure of impaired renal clearance, was entered in the primary model. Multivariable binary logistic regression was constrained to a maximum of 7 degrees of freedom, in line with the conventional rule of approximately 10 events per variable applied to the smaller outcome category (68 patients without hypoglycemia) [21]. Core covariates were selected a priori on pharmacological and physiological grounds: insulin use, sulfonylurea use, beta-blocker use, diabetic nephropathy, duration of diabetes (three categories) and site of care. Secondary behavioral and educational variables were added to the core model one at a time (core-plus-rotating strategy). Sensitivity analyses added age (four categories) and substituted CKD for nephropathy. Model calibration was assessed with the Hosmer–Lemeshow test. Results are reported as adjusted odds ratios (aOR) with 95% confidence intervals (CI); a two-sided p-value < 0.05 was considered statistically significant.

## Results

A total of 326 patients with T2DM aged 18 years and older participated in this study (Table 1). Slightly more than half were aged 61 years or older (52.8%; 39.6% aged 61–70 and 13.2% aged ≥71 years), and 54.6% were male. Mean weight and height were 85.0 kg (SD 18.9) and 167.4 cm (SD 9.3), respectively, consistent with the predominantly overweight-to-obese profile (37.1% overweight, 44.2% obese). Most participants resided in urban settings (55.2%) and had completed university-level education (51.2%); 30.7% were current or former smokers, and 75.8% reported no regular physical exercise.

**Table 1.** Sociodemographic and Anthropometric Characteristics of Study Participants (N=326)

| **Variable** | **n** | **%** |
|---|---|---|
| **Gender** |  |  |
| Female | 148 | 45.4 |
| Male | 178 | 54.6 |
| **Age (years)** |  |  |
| ≤ 50 | 45 | 13.8 |
| 51 – 60 | 109 | 33.4 |
| 61 – 70 | 129 | 39.6 |
| ≥ 71 | 43 | 13.2 |
| **Body Mass Index (kg/m²)** |  |  |
| < 25 | 61 | 18.7 |
| 25 – 29.9 | 121 | 37.1 |
| ≥ 30 | 144 | 44.2 |
| **Residence** |  |  |
| Rural | 146 | 44.8 |
| Urban | 180 | 55.2 |
| **Highest Educational Level** |  |  |
| Up to Primary School | 75 | 23.0 |
| Completed High School | 84 | 25.8 |
| Completed University, diploma or any higher education | 167 | 51.2 |
| **Exercise** |  |  |
| No | 247 | 75.8 |
| Yes | 79 | 24.2 |
| **Smoking Status** |  |  |
| Non-smoker | 226 | 69.3 |
| Smoker | 100 | 30.7 |
| **Recruitment Site** |  |  |
| KAUH | 197 | 60.4 |
| JUST PHC | 129 | 39.6 |

Most participants lived with family (96.9%) and had some form of health insurance (98.5%). By recruitment site, 197 participants (60.4%) were enrolled at KAUH and 129 (39.6%) at JUST PHC. Nearly two-fifths of participants were retired (39.3%). Reported monthly income was roughly split between less than 500 JD (44.5%) and 501–1000 JD (42.6%), with only 11.0% reporting income above 1000 JD.

More than half of patients (51.2%) had a diagnosis of T2DM for more than 10 years, while 26.1% had been diagnosed 5–10 years ago and 22.7% for less than 5 years. Only one third of the study population had an HbA1c within target range (≤ 7%), and most (56.4%) had this result recorded within the preceding 3 months. Overall, 119 patients (36.5%) had at least one diabetes-related microvascular complication, most commonly peripheral neuropathy (22.4%), followed by retinopathy (15.0%) and nephropathy (11.3%), while 9.8% had more than one such complication. Nearly three-quarters of patients (74.8%) had at least one comorbidity, most commonly hypertension (68.7%), followed by established cardiovascular disease (30.1%) and other non-cardiovascular comorbidities such as gout, hypothyroidism, or chronic liver disease (11.0%). Clinical characteristics of patients are shown in Table 2.

**Table 2.** Diabetes-Related Clinical Characteristics of Study Participants (N=326)

| **Variable** | **n** | **%** |
|---|---|---|
| **Years Diagnosed with Diabetes** |  |  |
| < 5 | 74 | 22.7 |
| 5 – 10 | 85 | 26.1 |
| > 10 | 167 | 51.2 |
| **HbA1c Level** |  |  |
| ≤ 7% | 110 | 33.7 |
| 7.1% – 8% | 82 | 25.2 |
| > 8% | 134 | 41.1 |
| **Diabetes-Related Microvascular Complications** |  |  |
| No complications | 207 | 63.5 |
| Single | 87 | 26.7 |
| Multiple | 32 | 9.8 |
| **General Comorbidities\*** |  |  |
| No comorbidity | 82 | 25.2 |
| Hypertension | 224 | 68.7 |
| Established CVD (IHD, CVA/TIA, PAD, HF) | 98 | 30.1 |
| Other non-CVD comorbidities (gout, hypothyroidism, liver disease) | 36 | 11.0 |

*\* Categories are not mutually exclusive; patients may have multiple comorbidities simultaneously. Consequently, the sum of percentages exceeds 100%.*

The prevalence of self-reported hypoglycemia during the past 6 months among patients with T2DM attending outpatient clinics was 79.1% (258/326). Among the 153 patients who provided detailed characteristics of their episodes, 5.2% required an emergency department visit or hospital admission, and 19.6% experienced at least one episode per week. Episodes most commonly occurred at midday (47.7%), followed by late night (28.8%), morning (24.8%), and evening (20.9%). Most patients (94.1%) managed episodes by self-treating with fast-acting carbohydrates or glucagon, while smaller proportions contacted a family member (8.5%) or visited a healthcare facility (3.3%); the large majority (88.2%) felt confident managing an episode independently. More than half of patients (53.6%) never reported their hypoglycemic episodes to a healthcare provider. Among the 113 patients who did not report at least some episodes, the most common reason was not believing the episode was serious (59.3%), followed by factors related to the healthcare provider (32.7%) and other patient-related factors (15.0%). Prevalence estimates and characteristics of reported and unreported hypoglycemia are shown in Table 3.

**Table 3.** Prevalence and Characteristics of Reported and Unreported Hypoglycemia.

| Variable | n | % |
|---|---|---|
| **In the past 6 months, have you had a blood glucose level below 70 mg/dL or experienced classical symptoms of hypoglycemia relieved by fast-acting carbohydrates?** |  |  |
| No | 68 | 20.9 |
| Yes | 258 | 79.1 |
| **Sub-group Analysis (n = 153)†** |  |  |
| **Hospitalization or ER Visit for Hypoglycemia (past 6 months)** |  |  |
| No | 145 | 94.8 |
| Yes | 8 | 5.2 |
| **Frequency of Hypoglycemic Episodes** |  |  |
| Rarely | 82 | 53.6 |
| Monthly | 41 | 26.8 |
| ≥ Weekly | 30 | 19.6 |
| **Time of Hypoglycemic Episodes‡** |  |  |
| Morning | 38 | 24.8 |
| Midday | 73 | 47.7 |
| Evening | 32 | 20.9 |
| Late Night (Nocturnal) | 44 | 28.8 |
| **Management of Hypoglycemic Episodes‡** |  |  |
| Self-treat with fast-acting carbohydrates or glucagon | 144 | 94.1 |
| Contact a family member | 13 | 8.5 |
| Visit a healthcare facility | 5 | 3.3 |
| **Confident Managing a Hypoglycemic Episode Independently§** |  |  |
| No | 18 | 11.8 |
| Yes | 135 | 88.2 |
| **Frequency of Reporting to a Healthcare Provider** |  |  |
| Never | 82 | 53.6 |
| Sometimes | 31 | 20.3 |
| Every time | 40 | 26.1 |
| **Reasons for Non-Reporting to a Healthcare Provider‡ (n = 113)** |  |  |
| "I thought it wasn't serious" | 67 | 59.3 |
| Factors related to healthcare provider | 37 | 32.7 |
| Other factors related to patient | 17 | 15.0 |

*† Analysis of event characteristics (hospitalization, frequency, timing, management, reporting) is restricted to the sub-group of patients who provided detailed event history (n=153); the remaining 105 patients reported hypoglycemia occurrence but did not specify characteristics. ‡ Categories are not mutually exclusive; patients may report more than one response. § Based on all 153 patients in the sub-group.*

Hypoglycemia prevalence was compared across sociodemographic, clinical, and treatment-related characteristics using Wilson score 95% confidence intervals and Pearson’s chi-square test (Table 4). Prevalence differed significantly by site of care, insulin use, and duration of diabetes. Patients attending JUST PHC had a higher prevalence of hypoglycemia than those attending KAUH (86.8% vs. 74.1%, p = 0.009). Patients taking insulin had a substantially higher prevalence than those not taking insulin (86.8% vs. 69.4%, p < 0.001), and prevalence increased with longer duration of diabetes (71.6% for <5 years, 75.3% for 5–10 years, and 84.4% for >10 years; p = 0.047). Prevalence did not differ significantly by gender, age group, sulfonylurea or β-blocker use, glycemic control, nephropathy, chronic kidney disease, BMI, education, or residence. These comparisons informed variable selection for the multivariable regression analysis presented in Table 6.

**Table 4.** Prevalence of Hypoglycemia by Selected Patient Characteristics.

| Variable | n | Prevalence, % (95% CI) | p-value |
|---|---|---|---|
| **Site\*** |  |  |  |
| KAUH | 197 | 74.1 (67.6–79.7) | 0.009 |
| JUST PHC | 129 | 86.8 (79.9–91.6) |  |
| **Gender** |  |  |  |
| Male | 178 | 80.9 (74.5–86.0) | 0.472 |
| Female | 148 | 77.0 (69.6–83.1) |  |
| **Age (years)** |  |  |  |
| ≤ 50 | 45 | 75.6 (61.3–85.8) | 0.728 |
| 51 – 60 | 109 | 78.0 (69.3–84.7) |  |
| 61 – 70 | 129 | 82.2 (74.7–87.8) |  |
| ≥ 71 | 43 | 76.7 (62.3–86.8) |  |
| **Insulin Use** |  |  |  |
| Yes | 182 | 86.8 (81.1–91.0) | < 0.001 |
| No | 144 | 69.4 (61.5–76.4) |  |
| **Sulfonylurea Use** |  |  |  |
| Yes | 196 | 82.1 (76.2–86.9) | 0.134 |
| No | 130 | 74.6 (66.5–81.3) |  |
| **β-Blocker Use** |  |  |  |
| Yes | 126 | 74.6 (66.4–81.4) | 0.144 |
| No | 200 | 82.0 (76.1–86.7) |  |
| **HbA1c Level** |  |  |  |
| Controlled (≤ 7%) | 110 | 73.6 (64.7–81.0) | 0.109 |
| Uncontrolled (> 7%) | 216 | 81.9 (76.3–86.5) |  |
| **Duration of Diabetes** |  |  |  |
| < 5 years | 74 | 71.6 (60.5–80.6) | 0.047 |
| 5 – 10 years | 85 | 75.3 (65.2–83.2) |  |
| > 10 years | 167 | 84.4 (78.2–89.1) |  |
| **Nephropathy** |  |  |  |
| Yes | 37 | 91.9 (78.7–97.2) | 0.070 |
| No | 289 | 77.5 (72.4–81.9) |  |
| **Chronic Kidney Disease** |  |  |  |
| Yes | 22 | 90.9 (72.2–97.5) | 0.256 |
| No | 304 | 78.3 (73.3–82.6) |  |
| **Body Mass Index (kg/m²)** |  |  |  |
| < 25 | 61 | 73.8 (61.6–83.2) | 0.518 |
| 25 – 29.9 | 121 | 80.2 (72.2–86.3) |  |
| ≥ 30 | 144 | 80.6 (73.3–86.2) |  |
| **Highest Educational Level** |  |  |  |
| Up to Primary School | 75 | 70.7 (59.6–79.8) | 0.059 |
| Completed High School | 84 | 77.4 (67.4–85.0) |  |
| Completed University, diploma or any higher education | 167 | 83.8 (77.5–88.6) |  |
| **Residence** |  |  |  |
| Urban | 180 | 80.0 (73.6–85.2) | 0.774 |
| Rural | 146 | 78.1 (70.7–84.0) |  |

*CI: confidence interval, calculated using the Wilson score method. P-values from Pearson's chi-square test (continuity-corrected for 2×2 comparisons).*

Most patients (88.7%) reported monitoring their blood glucose (Table 5), most commonly weekly (40.6%) or daily (32.6%) among those with a recorded frequency (n=288), and self-monitoring was the predominant method (75.5%), followed by a family member (11.0%) or a health professional (13.5%). Only about one-third of patients (32.8%) had heard of continuous glucose monitoring (CGM) technology. While most patients (71.5%) knew the signs and symptoms of hypoglycemia, the large majority (73.9%) were unaware that hypoglycemia can occur without symptoms. Most patients (68.7%) had never discussed hypoglycemia risk with their physician, and 58.9% had not received written or verbal instructions on its management. About half of patients (50.6%) never carried sugar-containing food or an emergency kit for hypoglycemia, and 41.7% visited their healthcare provider fewer than three times per year. Independently, 49.7% of patients reported skipping at least one meal during the day, and 89.3% reported practicing fasting for religious or health reasons. One in five patients (20.2%) experienced anxiety related to hypoglycemia, and among these, adherence to antihyperglycemic drugs was affected in nearly half (48.5%, n=66). Diabetes monitoring, knowledge, and risk behaviors are summarized in Table 5.

**Table 5.** Diabetes Monitoring, Knowledge, and Risk Behaviors.

| Variable | n | % |
|---|---|---|
| **Blood Glucose Monitoring** |  |  |
| No | 37 | 11.3 |
| Yes | 289 | 88.7 |
| **Frequency of Blood Glucose Monitoring (n = 288)\*** |  |  |
| Once a Month or Less | 77 | 26.7 |
| Weekly | 117 | 40.6 |
| Daily | 94 | 32.6 |
| **Primary Blood Glucose Monitor** |  |  |
| Self | 246 | 75.5 |
| Family | 36 | 11.0 |
| Health Professional | 44 | 13.5 |
| **Awareness of Hypoglycemia-Related Signs and Symptoms** |  |  |
| No | 93 | 28.5 |
| Yes | 233 | 71.5 |
| **Awareness of Asymptomatic Hypoglycemia Occurrence** |  |  |
| No | 241 | 73.9 |
| Yes | 85 | 26.1 |
| **Physician Discussed Hypoglycemia Risk and Management** |  |  |
| No | 224 | 68.7 |
| Yes | 102 | 31.3 |
| **Physician Provided Written or Verbal Instructions on Managing Hypoglycemia** |  |  |
| No | 192 | 58.9 |
| Yes | 134 | 41.1 |
| **Carrying Sugar-Containing Food or an Emergency Kit for Hypoglycemia** |  |  |
| Never | 165 | 50.6 |
| Sometimes | 41 | 12.6 |
| Always | 120 | 36.8 |
| **Skipping Meals During the Day** |  |  |
| No | 164 | 50.3 |
| Yes | 162 | 49.7 |
| **Practice Fasting** |  |  |
| No | 35 | 10.7 |
| Yes | 291 | 89.3 |
| **Healthcare Provider Visit Frequency** |  |  |
| Less than 3 Times a Year | 136 | 41.7 |
| Every 3 Months | 120 | 36.8 |
| Every Month | 70 | 21.5 |
| **Anxiety Related to Hypoglycemia** |  |  |
| No | 260 | 79.8 |
| Yes | 66 | 20.2 |
| **Adherence to Antihyperglycemic Drugs Affected by Hypoglycemia Anxiety (n = 66)** |  |  |
| No | 34 | 51.5 |
| Yes | 32 | 48.5 |

*\* Percentages calculated among the 288 of 289 monitoring patients with a recorded frequency (one record missing).*

Multivariable logistic regression (Table 6) identified factors independently associated with hypoglycemia. In the core model (Hosmer–Lemeshow p = 0.223), insulin use (aOR 3.27, 95% CI 1.75–6.10), sulfonylurea use (aOR 2.14, 1.18–3.87) and attending JUST PHC rather than KAUH (aOR 2.76, 1.42– 5.36) were independently associated with higher odds of hypoglycemia. Diabetic nephropathy was also associated with higher odds, with an imprecise estimate (aOR 3.76, 1.05–13.48; 37 patients). Beta-blocker use was associated with lower odds (aOR 0.52, 0.28–0.95). Duration of diabetes was not independently associated with hypoglycemia. When added individually to the core model, skipping meals was independently associated with hypoglycemia (aOR 2.44, 1.33–4.49), whereas education (university vs primary: aOR 2.05, 1.00–4.22), gender, age, exercise, fasting, incorrect dosing, awareness of asymptomatic hypoglycemia and physician discussion were not. Additional adjustment for age in four categories did not materially change the estimates (likelihood-ratio test p = 0.532), and substituting CKD for nephropathy gave similar results for the other covariates.

**Table 6.** Multivariable Logistic Regression of Factors Associated with Self-Reported Hypoglycemia (N=326; 258 events)

| **Variable** | **aOR** | **95% CI** | **p** |
|---|---|---|---|
| **Core model (7 df)** |  |  |  |
| Insulin use (yes vs no) | 3.27 | 1.75–6.10 | < 0.001 |
| Sulfonylurea use (yes vs no) | 2.14 | 1.18–3.87 | 0.012 |
| Beta-blocker use (yes vs no) | 0.52 | 0.28–0.95 | 0.033 |
| Diabetic nephropathy (yes vs no) | 3.76 | 1.05–13.48 | 0.042 |
| Duration 5–10 vs < 5 years | 0.81 | 0.38–1.75 | 0.596 |
| Duration > 10 vs < 5 years | 1.47 | 0.72–3.00 | 0.289 |
| Site: JUST PHC vs KAUH | 2.76 | 1.42–5.36 | 0.003 |
| **Core + one variable at a time\*** |  |  |  |
| Skipping meals (yes vs no) | 2.44 | 1.33–4.49 | 0.004 |
| Education: high school vs primary | 1.33 | 0.62–2.86 | 0.471 |
| Education: university vs primary | 2.05 | 1.00–4.22 | 0.051 |
| Incorrect medication dosage (ever) | 1.25 | 0.49–3.22 | 0.641 |
| Aware hypoglycemia can be asymptomatic | 1.45 | 0.73–2.89 | 0.285 |
| Physician discussed hypoglycemia risk | 1.69 | 0.84–3.41 | 0.139 |
| Gender (male vs female) | 0.93 | 0.52–1.68 | 0.815 |
| Age > 60 vs ≤ 60 years | 1.34 | 0.74–2.43 | 0.329 |
| Regular exercise (yes vs no) | 0.81 | 0.40–1.65 | 0.564 |
| Practicing fasting (yes vs no) | 1.18 | 0.45–3.06 | 0.740 |
| **Sensitivity analyses** |  |  |  |
| (a) Core + age, four categories (10 df)†: 51–60 vs ≤ 50 | 1.26 | 0.51–3.09 | 0.619 |
| 61–70 vs ≤ 50 | 1.79 | 0.72–4.45 | 0.213 |
| ≥ 71 vs ≤ 50 | 1.11 | 0.37–3.37 | 0.854 |
| Nephropathy in this model | 3.96 | 1.08–14.53 | 0.038 |
| (b) CKD substituted for nephropathy (7 df)‡ | 3.50 | 0.74–16.55 | 0.113 |

*aOR, adjusted odds ratio. Core model Hosmer–Lemeshow p = 0.223. \* Each row is from a separate model: the core covariates plus the single listed variable; Hosmer–Lemeshow p ≥ 0.058 in all. † Likelihood-ratio test for age (3 df) p = 0.532; Hosmer–Lemeshow p = 0.369. This model uses 10 df, exceeding the events-per-variable limit, and is reported as a sensitivity analysis only; age as a single ordinal term gave aOR 1.12 per category (0.80–1.58), p = 0.505. Other covariates in model (a): insulin 3.40, sulfonylurea 2.23, beta-blocker 0.49, JUST PHC 2.70. ‡ Other covariates in model (b): insulin 3.26, sulfonylurea 2.11, beta-blocker 0.53, JUST PHC 2.73. Hypoglycemia-related anxiety and carrying sugar were not modelled as risk factors because they are likely consequences of hypoglycemia.*

## Discussion

### Principal findings

In this cross-sectional study of 326 adults with type 2 diabetes mellitus (T2DM) attending outpatient clinics in northern Jordan, four in five patients (79.1%) reported at least one hypoglycemic episode in the preceding six months. Awareness of hypoglycemia was incomplete: although most participants recognized its symptoms, fewer than three in ten knew that hypoglycemia can occur without symptoms, over half had never reported an episode to a healthcare provider, and half did not routinely carry a source of fast-acting sugar. In the multivariable model, insulin use, sulfonylurea use, diabetic nephropathy and attendance at the primary healthcare clinic (versus the tertiary hospital) were independently associated with higher odds of hypoglycemia, whereas beta-blocker use was associated with lower odds. Age, diabetes duration and sex were not independently associated with the outcome.

### Prevalence of hypoglycemia

The prevalence we observed is higher than most published estimates, including those for patient-reported hypoglycemia. In a review of the epidemiology of hypoglycemia in T2DM, 46–58% of patients not receiving insulin reported any hypoglycemic symptoms over six months, and 37–64% of insulin-treated patients did so over twelve months [22]. Earlier Jordanian outpatient studies reported lower figures, including 46.6% over three months among 1,055 patients attending the National Center for Diabetes, Endocrinology and Genetics in Amman, where hypoglycemia was defined by symptoms alone [18], and 21.7% among older outpatients at King Abdullah University Hospital [19]. Several features of our design plausibly explain this difference. First, our case definition was deliberately sensitive, combining a measured glucose below 70 mg/dL with symptoms relieved by fast-acting carbohydrate, so it captures non-severe and biochemically unconfirmed events that stricter definitions exclude [4,20]. Second, the six-month recall window cumulates events over a longer period than the one-week windows used elsewhere [23]. Third, the sample was drawn from clinic attenders with a high-risk treatment profile: 55.8% were using insulin, 60.1% a sulfonylurea, and 51.2% had lived with diabetes for more than ten years. Fourth, symptom recall may be inflated by non-hypoglycemic causes of similar symptoms, as symptomatic thresholds are known to vary with glycemic control and prior exposure [14–16]. The estimate should therefore be read as the burden of self-perceived hypoglycemia among high-risk clinic attenders, not as a population incidence, and comparisons with studies that used different definitions and recall periods should be made cautiously.

### Differences between the two clinical settings

Patients attending JUST PHC had 2.76-fold higher adjusted odds of hypoglycemia than those attending KAUH (95% CI 1.42–5.36). This association persisted after adjustment for treatment, nephropathy and diabetes duration, so it is not explained by differences in those factors. Because the sites differ in case mix and care processes, site is best read as a marker of those differences, not of the quality of care. Established risk factors for hypoglycemia in type 2 diabetes are patient- and regimen-related [22].

In post hoc exploratory analyses, glycemic control differed by site: 41.9% of PHC patients reported an HbA1c ≤7% versus 28.4% at KAUH (p = 0.017). The PHC advantage in control was attenuated after adjustment for age, diabetes duration, insulin, sulfonylurea use, nephropathy and education (aOR for HbA1c >7% 0.62, 95% CI 0.37–1.04). The site difference in hypoglycemia was concentrated among patients with an HbA1c ≤7% (87.0% vs 60.7%; aOR 6.11, 95% CI 2.02–18.47) and was not evident among those above 7% (86.7% vs 79.4%; aOR 1.52, 95% CI 0.63–3.64). Hypoglycemia at the PHC was therefore uniformly frequent irrespective of glycemic control, whereas at KAUH it was less frequent among well-controlled patients. The site-by-control interaction was not statistically significant (p = 0.16) and the estimates are imprecise, so these findings are hypothesis-generating. One plausible interpretation is that glucose-lowering therapy was not de-intensified in patients who had reached tight targets. Such potential overtreatment is documented across practice settings, including primary care, in US adults aged ≥75 years [24].

Other exploratory site differences were consistent with this picture. PHC patients were less likely to have received written or verbal instructions on managing hypoglycemia (31.0% vs 47.7%, p = 0.004), less likely to take beta-blockers (26.4% vs 46.7%, p < 0.001), and more likely to practice fasting (96.1% vs 84.8%, p = 0.002). Limited hypoglycemia counseling in primary care visits has been described elsewhere [25]. Because beta-blocker use was more common at KAUH and was itself inversely associated with hypoglycemia, the site and beta-blocker terms are partly entangled. Residual confounding (for example, specific sulfonylurea or insulin regimens, renal function and diet) and site differences in symptom perception or recall are equally plausible.

These findings should not be read as evidence of inferior primary care. They identify primary care as a priority setting for hypoglycemia prevention, including structured patient education and periodic review of treatment intensity against individualized targets.

### Reporting and communication with healthcare providers

More than half of participants (53.6%) had never reported a hypoglycemic episode to a healthcare provider, and 59.3% of those who did not report considered the episode not serious enough. Only 31.3% recalled a physician discussing hypoglycemia risk with them. Under-reporting is well described: in a seven-country European survey, 50–59% of insulin-treated patients with T2DM rarely or never told their physician about hypoglycemic events, and 26% were not asked about hypoglycemia at routine appointments [23]. Barriers include competing priorities during consultations, limited recall and logging of events, and fear of blame [26]. In our setting, where 41.7% of patients attend a provider fewer than three times a year, a missed opportunity to ask about hypoglycemia has an outsized effect, and the clinical burden recorded in medical files is likely to underestimate the true burden.

### Awareness of hypoglycemia

Although 71.5% of participants correctly identified hypoglycemic symptoms, 73.9% were unaware that hypoglycemia can be asymptomatic. This gap matters because impaired awareness of hypoglycemia is common in T2DM, with a pooled prevalence of about 22% in one meta-analysis [27]; among people with diabetes, prevalence varied from roughly 23% to 59% depending on the instrument in another [28], and impaired awareness increases the risk of severe events. A study of insulin-treated patients in Saudi Arabia found that poor diabetes knowledge was among the main risk factors for unawareness [29]. We did not use a validated instrument such as the Clarke or Gold score, so our knowledge item measures awareness that asymptomatic hypoglycemia exists, not whether individual patients have impaired awareness. Self-management behaviors also reflected this gap: only 32.6% of those who monitored did so daily and two-thirds tested weekly or less, only 32.8% had heard of continuous glucose monitoring, and 50.6% never carried sugar. Because 64 of 66 patients who reported anxiety about hypoglycemia had experienced an episode, and carrying sugar may follow rather than precede hypoglycemia, we did not enter these variables in the multivariable model; the temporal ordering cannot be resolved in a cross-sectional design.

### Factors associated with hypoglycemia

The associations with insulin (adjusted OR 3.27) and sulfonylurea use (adjusted OR 2.14) are consistent with established pharmacology and clinical guidance, which identify these agents as the principal drug-related causes of hypoglycemia in T2DM [2,3,20,22]. Both remained independent of each other, supporting individualized review of sulfonylurea use, particularly in patients with other risk factors.

Diabetic nephropathy was associated with 3.76-fold higher odds, although with a wide confidence interval (1.05–13.48) reflecting the small number of affected patients. Diabetic nephropathy was also significantly associated with hypoglycemia among patients at the National Center for Diabetes, Endocrinology and Genetics [18]. Reduced renal clearance of insulin and sulfonylureas and impaired renal gluconeogenesis are recognized mechanisms, and the incidence of hypoglycemia in chronic kidney disease is roughly twice that in people without CKD among patients with diabetes [30]. Because 21 of the 22 patients with CKD also had nephropathy, the two terms overlap; when CKD was substituted for nephropathy the estimate was similar in magnitude but not statistically significant (adjusted OR 3.50, 95% CI 0.74–16.55), and this finding should be treated as hypothesis-generating.

When added individually to the core model, skipping meals was independently associated with hypoglycemia (aOR 2.44, 95% CI 1.33–4.49), as expected; it was not included in the core model itself because of the events-per-variable constraint, so this estimate is adjusted for the core covariates but not for the other variables examined one at a time. Fasting was practiced by 89.3% of participants and was not associated with hypoglycemia when added to the core model (aOR 1.18), but with so little variation the study had little power to detect an effect. Structured pre-Ramadan risk stratification and education are recommended by international guidance to reduce hypoglycemia in fasting patients [31], and our data on instruction by providers suggest this is an area for improvement. Higher education showed a borderline association (university education, added to the core model: aOR 2.05, 95% CI 1.00–4.22, p = 0.051); better-educated patients may be more attentive to, or more willing to report, symptoms, so this may reflect detection rather than risk. Neither age nor diabetes duration was independently associated with hypoglycemia, including when age was modelled in four categories (likelihood-ratio p = 0.532), indicating that hypoglycemia risk in this sample is driven more by treatment and comorbidity than by chronological age or time since diagnosis.

### Beta-blocker use and lower odds of hypoglycemia

Beta-blocker use was associated with 48% lower odds of hypoglycemia (adjusted OR 0.52, 95% CI 0.28– 0.95). This is unexpected, since beta-blockers can mask adrenergic warning symptoms and have been associated with higher rates of severe hypoglycemia, including among sulfonylurea users and in the ACCORD trial population [32,33]. Our outcome relied on recognized symptoms or measured glucose, so masking of adrenergic symptoms could have led to fewer episodes being perceived and reported, which would produce a spurious inverse association. Other possible explanations include confounding by indication (patients prescribed beta-blockers may be older with cardiovascular disease and treated less intensively) and chance, given the borderline p-value (0.033) and wide interval. The finding should not be interpreted as protective and does not alter the advice that patients taking beta-blockers with insulin or sulfonylureas need particular attention to hypoglycemia monitoring.

### Strengths and limitations

Strengths include recruitment from two clinical settings in northern Jordan, a pre-specified analysis plan with an events-per-variable limit to reduce overfitting [21], adequate model calibration (Hosmer–Lemeshow p = 0.223), transparent reporting of sensitivity analyses, and the coverage of both clinical correlates and patient awareness and behavior, an area with little published data from Jordan.

Several limitations should be considered. First, the cross-sectional design precludes causal inference and temporal ordering, particularly for anxiety, carrying sugar, and monitoring behaviors. Second, hypoglycemia was self-reported over six months, so recall bias and misclassification are likely, and the definition did not require biochemical confirmation or distinguish severity; we cannot report the prevalence of severe hypoglycemia. Third, the sample was a convenience sample of clinic attenders, which limits generalizability to patients who do not attend or are less engaged with care, and it was drawn from two settings in one region of Jordan. Fourth, with only 68 patients without hypoglycemia, the multivariable model was restricted to seven degrees of freedom, so important covariates (for example, HbA1c, specific drug agents, renal function, and diet) could not be adjusted for simultaneously, and some confidence intervals are wide. Fifth, awareness was assessed with non-validated items, and the sensitivity of the outcome to symptom-based criteria may overstate true hypoglycemia. Sixth, the site variable was derived from the medical file number, and some subgroup items were completed by fewer participants, so site differences should be interpreted cautiously. Finally, patients with type 1 diabetes were excluded and so findings apply only to T2DM.

### Implications for practice and research

These findings support several practical measures in Jordanian outpatient diabetes care: routine, structured questioning about hypoglycemia at every visit; patient education that explicitly addresses asymptomatic hypoglycemia, the signs that may be masked by beta-blockers, and the need to carry fast-acting carbohydrate; review of sulfonylurea use and insulin dosing in patients with nephropathy or other risk factors; and pre-fasting counseling before Ramadan. Wider access to glucose monitoring, including continuous monitoring where feasible, should be considered for those at highest risk [2,20]. Future studies should use validated awareness instruments, confirm events with glucose records or sensor data, measure severity, and follow patients prospectively to clarify the roles of treatment intensity, renal function and beta-blockers.

## Conclusion

Self-reported hypoglycemia was highly prevalent among patients with T2DM attending outpatient clinics in northern Jordan, and awareness of its asymptomatic form and of the need to report it was limited. Insulin, sulfonylureas, diabetic nephropathy and attendance at primary care clinics were independently associated with hypoglycemia. Targeted education, routine screening for hypoglycemia at clinic visits, and careful review of high-risk therapy are warranted.

## Author contributions

Adi Khasawneh: Conceptualization, Methodology, Supervision, Writing – review & editing. Taima’a Kefah Al-Mohammad: Data curation, Formal analysis, Writing – original draft. Karam Mohamad Aldeeb: Data curation, Investigation, Writing – original draft. Sohaib Fawaz Shaheen: Data curation, Investigation, Writing – original draft. Razan Yaseen: Data curation, Investigation, Writing – original draft. Sara Ali Nassar: Data curation, Investigation. Mohammad Emad Melhem: Data curation, Investigation. Abdel-Hameed Al-Mistarehi: Formal analysis, Methodology, Writing – review & editing. Abdalrahman Abu Qutaish: Formal analysis. Heba Okour: Formal analysis, Writing – review & editing. Khalid A. Kheirallah: Conceptualization, Funding acquisition, Methodology, Project administration, Supervision, Writing – review & editing.

## Acknowledgments

The authors wish to thank the physicians and clinic staff at King Abdullah University Hospital and the JUST Primary Healthcare Centre for their cooperation in facilitating data collection, and all participants who volunteered their time for this study.

## Data Availability

The data underlying this study are not publicly deposited due to privacy considerations related to patient health information. The datasets used and analysed during the current study are available from the corresponding author (Khalid A. Kheirallah) upon reasonable request and following institutional approval.

## Supporting Information

S1 STROBE Checklist. STROBE checklist for cross-sectional studies.

